# Evaluating elexacaftor/tezacaftor/ivacaftor (ETI; Trikafta™) for treatment of patients with non-cystic fibrosis bronchiectasis (NCFB): a clinical study protocol

**DOI:** 10.1101/2024.07.19.24310604

**Authors:** Colin E. Swenson, William R. Hunt, Candela Manfredi, Diana J. Beltran, Jeong S. Hong, Brian R. Davis, Shingo Suzuki, Cristina Barillá, Andras Rab, Cynthia Chico, Joy Dangerfield, Ashleigh Streby, Elizabeth M. Cox, Arlene Stecenko, Adrianna Westbrook, Rebecca Kapolka, Eric J. Sorscher

## Abstract

Non-cystic fibrosis bronchiectasis (NCFB) is a disease characterized by abnormal dilatation of the airways, airflow obstruction, persistent cough, excessive sputum production and recurrent lung infections. NCFB exhibits clinical and pathological manifestations similar to key features of cystic fibrosis (CF) lung disease. In CF, pathogenesis results from dysfunction of the cystic fibrosis transmembrane conductance regulator (CFTR), and diagnosis is made by demonstrating elevated sweat chloride concentrations (typically ≥60 mEq/L), two CFTR mutations known to be causal, multi-organ tissue injury, or combination(s) of these findings. Based on a considerable body of evidence, we believe many patients with NCFB have disease likely to benefit from drugs such as elexacaftor/tezacaftor/ivacaftor (ETI) that activate CFTR-dependent ion transport. ETI is currently prescribed solely for treatment of CF, and has not been adequately tested or proposed for patients with NCFB, many of whom exhibit decreased CFTR function. Accordingly, we are conducting a clinical trial of ETI in subjects carrying a diagnosis of NCFB. Participants will exhibit one disease-causing CFTR mutation and/or sweat chloride measurements of 30-59 mEq/L. Cutaneous punch biopsy or blood samples will be obtained for iPS cell differentiation into airway epithelial monolayers – which will then be tested for response to ETI. Each patient will be given CFTR modulator treatment for approximately four weeks, with monitoring of clinical endpoints that include FEV_1_, sweat chloride, quality of life questionnaire, and weight. The study will evaluate response of patients with NCFB to ETI, and test usefulness of iPSC-derived airway epithelial monolayers as a novel *in vitro* technology for predicting clinical benefit.

**FUNDING SOURCE:** Marcus Foundation, Inc

**“Main Study” Summary:** 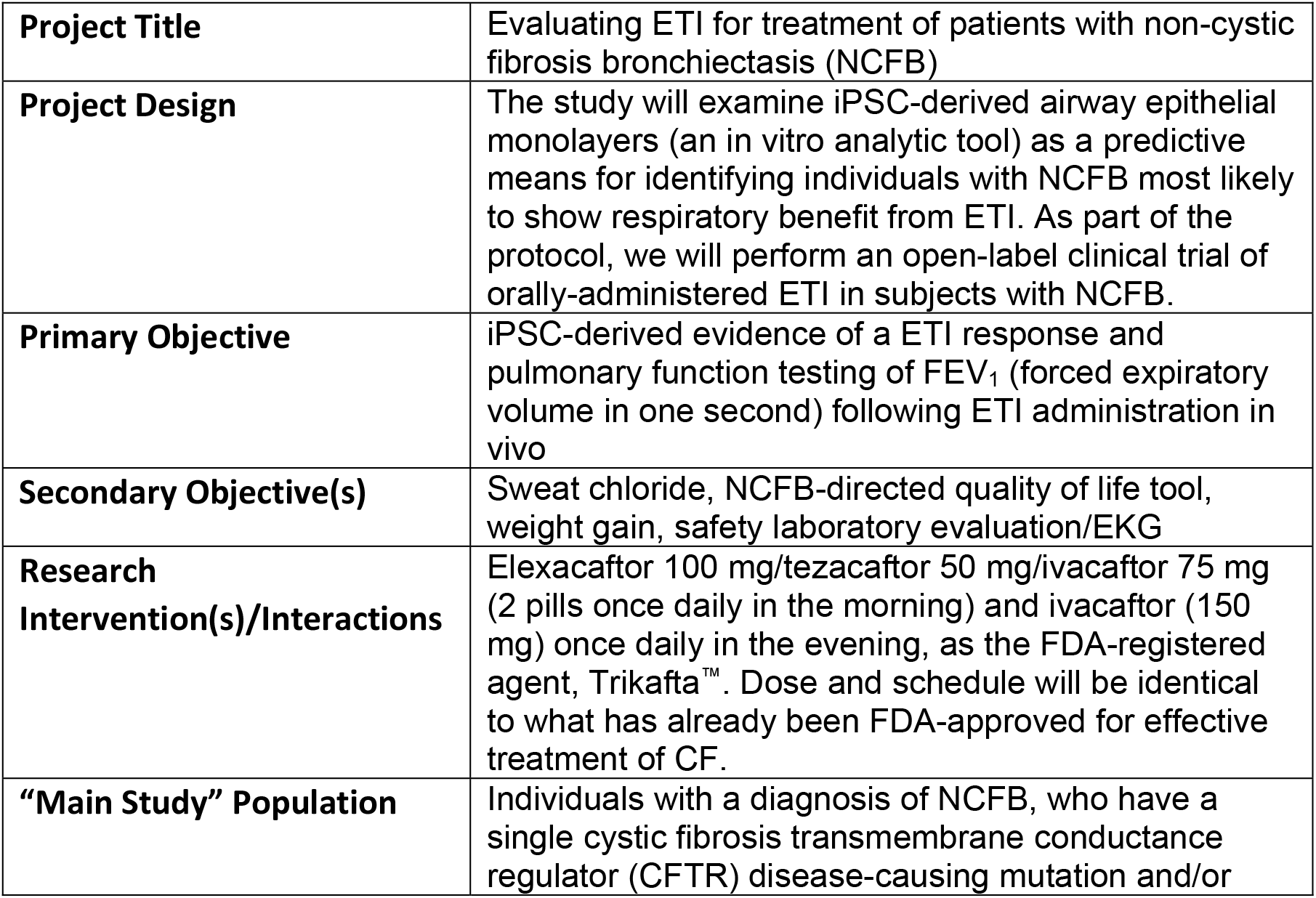

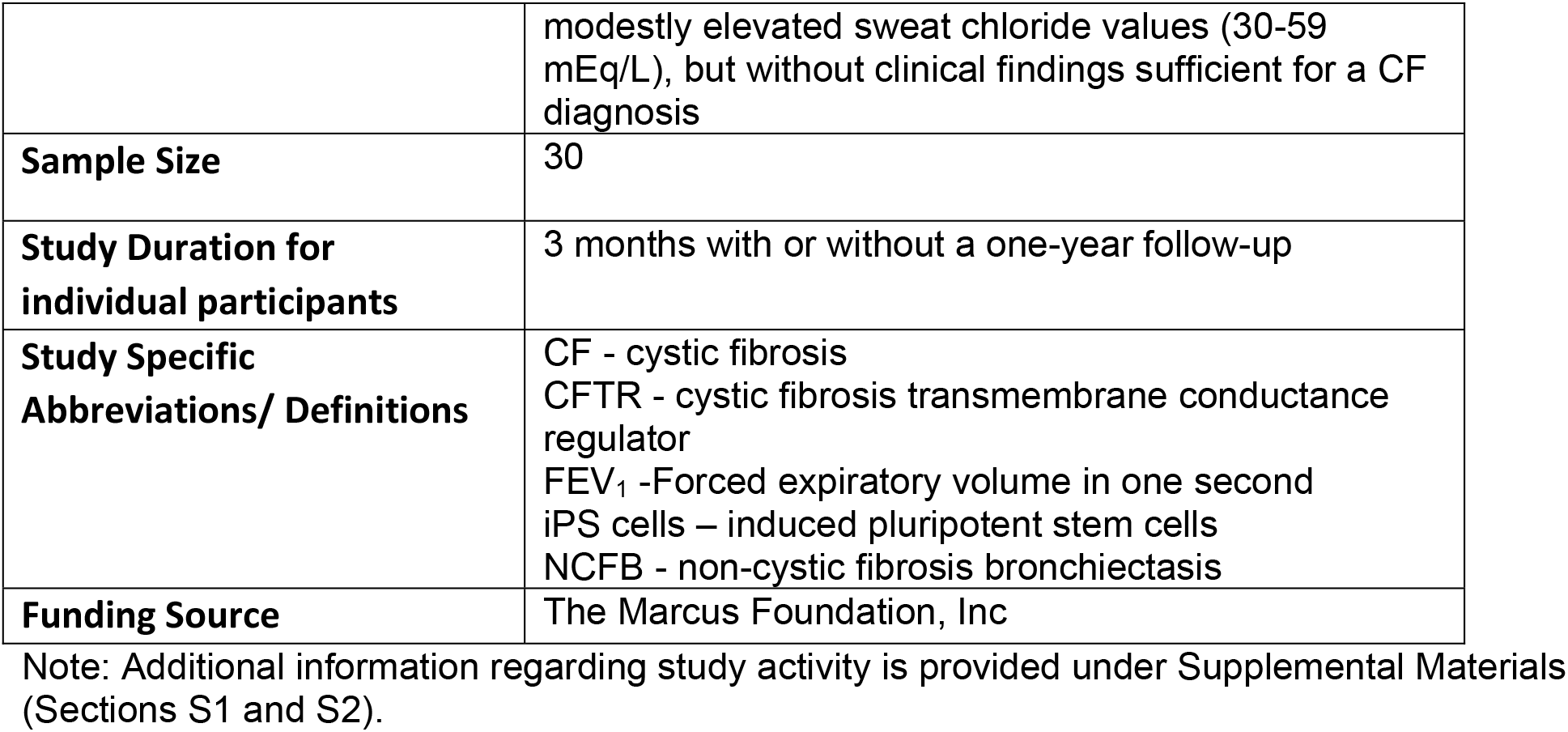

**Objectives:** *‘Lead in’ study: Incidence of diminished CFTR activity among patients with NCFB:* In order to gain information regarding numbers of individuals with NCFB who may be eligible for our ‘main’ study, patients at Emory followed with bronchiectasis (who do not have clinical criteria sufficient for a diagnosis of cystic fibrosis) will be asked to consider participating in a ‘lead in’ study that will determine the subject’s CFTR genotype and sweat chloride level. A separate consent form will be utilized for the ‘lead in’ study. Subjects who exhibit a single CF-causing mutation in CFTR and/or sweat chloride 30-59 mEq/L will be approached about their interest in reviewing the consent form for the ‘main’ study (“iPSC derivation and *in vivo* ETI treatment for 4 weeks”; see following section). Up to 200 subjects will be included in the ‘lead-in’ study.

*‘Main’ study: iPSC derivation and *in vivo* ETI treatment for 4 weeks:* We propose a clinical trial of subjects with a diagnosis of NCFB. Participants will exhibit one disease-causing CFTR mutation and/or sweat chloride measurements of 30-59 mEq/L. Each patient will be given ETI for approximately four weeks. We will monitor clinical endpoints that include FEV_1_, sweat chloride, quality of life questionnaire, and weight. We will also collect cutaneous punch biopsy material or blood samples from subjects so iPS cells can be differentiated into airway epithelial monolayers and tested for response to ETI.

## Background

Non-cystic fibrosis bronchiectasis is a clinical syndrome characterized by abnormal dilatation of the airways, airflow obstruction, persistent cough, excessive sputum production and recurrent lung infections. In terms of pathophysiology, airway dilatation and other features are associated with impaired mucociliary clearance and failure to adequately clear bacteria and mucus secretions from the airways. These events contribute to persistent infection, inflammation, and further progressive airway damage, leading to diminished lung function, with the possibility of respiratory failure and death. The pathogenesis of non-CF bronchiectasis is complex, poorly understood, and is likely to vary depending on the underlying etiology and important modifying factors ^1-3^.

NCFB is clinically and pathologically similar to certain features of cystic fibrosis lung disease. For example, like CF, NCFB pulmonary injury is characterized by pronounced bronchiectasis, airway architectural damage, mucus accumulation, chronic lung infection and persistent neutrophilic infiltrates^2-6^. In CF, disease is caused by generalized dysfunction of the cystic fibrosis transmembrane conductance regulator, and diagnosis is made by demonstrating elevated sweat chloride concentrations (typically ≥60 mEq/L), two CFTR mutations shown to be pathogenic, multi-organ disease, or a combination of these findings. By convention, sweat chloride levels in NCFB are <60 mEq/L. NCFB is therefore often considered a diagnosis of exclusion – if criteria are inadequate to establish a diagnosis of “cystic fibrosis,” NCFB may instead be entertained. Patients with NCFB are not approved for ETI, and do not have access to the drug (see also Supplementary Materials, S3). Based on a considerable body of evidence, we believe a substantial number of patients with NCFB have disease likely to exhibit clinical benefit from drugs such as ETI that activate CFTR-dependent ion transport. Such a notion has not been adequately tested or proposed previously.

In a subset of individuals with NCFB, pathogenesis is likely due – at least in part – to CFTR deficiency, originating from inherited factors (i.e., asymptomatic CF carrier status, which occurs in approximately 1 of 30 Caucasians in the US and leads to ∼50% decrease in CFTR mRNA and protein), acquired factors (e.g., due to chronic airway inflammation, infection, hypoxia, or toxin exposure), or a combination of these. For example, in patients with chronic airway infection and inflammation, or past toxin (e.g. nicotine) exposure, CFTR activity has been suggested to be negatively impacted – resulting in defects in anion secretion, airway surface liquid depletion and blunted mucociliary transport^7,8^. Furthermore, CFTR mutation carriers, with an estimated half the level of functional CFTR compared to the general population, are at increased risk for a range of adverse conditions, including NCFB^9-12^. Moreover, in a study of 122 individuals with NCFB and normal sweat chloride concentrations, 22 (18%) were found to have one CFTR mutation and abnormal CFTR function in respiratory track epithelium that was intermediate between healthy controls and those with classical CF^13^.

### Considerations regarding NCFB

The incidence of NCFB is increasing worldwide. Previously classified as a rare or orphan disease, the diagnosis has increased by 40% in the past several years ^1^ and the number of adults with bronchiectasis in the United States is estimated at 350,000 - 500,000 ^14^; i.e., much higher than the number of adults with CF (which amount to 30,000-40,000). Unfortunately, there are no curative therapies or medications specifically approved to address fundamental mechanisms that underlie NCFB.

ETI is approved for patients with CF carrying at least one copy of the common F508del variant or a number of other CFTR abnormalities. Trikafta^™^ is a combination of three CF drugs, elexacaftor, ivacaftor, and tezacaftor (ETI), that helps CFTR proteins work more effectively. Patients with common forms of CF typically exhibit robust pulmonary benefit from ETI within several days of initiating treatment^6,15,16^.

ETI is not prescribed or approved for NCFB and has not been considered of benefit for patients with NCFB. We hypothesize that a drug such as Trikafta^™^, which markedly activates both wild type and mutant CFTR *in vivo*, will enhance mucociliary clearance and should improve respiratory function (FEV_1_) in a subset of patients with NCFB. Even if one in ten patients with NCFB shows significant improvement in FEV_1_ following ETI, this would constitute a major discovery and provide new hope for tens of thousands of individuals in the US with an otherwise untreatable and lethal lung disease.

### Emerging approaches to identifying patients with NCFB most likely to exhibit clinical response to ETI

The nature and extent of *in vitro* model systems that predict pulmonary responsiveness to ETI are being evaluated by several laboratories, including our own. Reprogramming of adult somatic cells into iPSCs is a powerful approach that holds promise for regenerative and personalized medicine. This system presents potential advantages over classical *in vitro* cell-based and other models, and has been useful for generating efficient, high-performance tools for clinical/translational research and drug development^17^. From a translational perspective, the use of patient-derived specimens may allow a comparative test of drug efficacies *in vitro* and therapeutic responses at an individual level, as recently evaluated by a number of investigators, including those participating in the current project ^18-25^.

#### Summary

We believe NCFB in many individuals will respond to stimulation of CFTR function in the airway, including patients with sweat chloride values that are normal or only mildly elevated (i.e., indicating diminished CFTR activity). The present study will comprise an open-label, one center trial of orally administered elexacaftor, tezacaftor and ivacaftor (ETI) that will enroll 30 patients with NCFB. Study subjects will have one known CFTR mutation and/or mildly elevated sweat chloride measurements (i.e., 30-59 mEq/L; Figure 1 summarizes conventional trial endpoints). Patients with a diagnosis of CF will be excluded (see below). A detailed schedule of events and other information is provided under “Human Subject Involvement” and Supplemental Materials S1. The study will specifically and prospectively test iPS cells taken from patients with NCFB to determine *in vitro* thresholds for predicting CFTR rescue *in vivo*. Using iPS cells differentiated to exhibit a respiratory epithelial phenotype, we will determine whether the emerging technology can be used to predict FEV_1_ response among individuals with NCFB who receive ETI.

**Figure 1.**
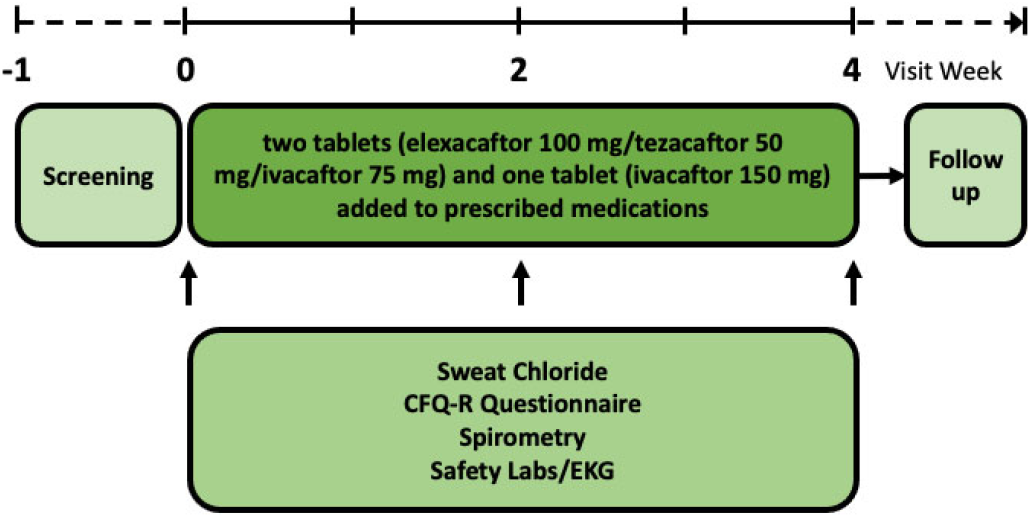
Summary of trial design.

### “Main Study” Endpoints

- Primary Endpoints: iPSC-derived evidence of a ETI response and pulmonary function testing of FEV_1_ (forced expiratory volume in one second) following ETI administration *in vivo*
- Secondary Endpoints: sweat chloride, NCFB-directed quality of life tool, weight gain, safety laboratory evaluation/EKG

| OBJECTIVES | ENDPOINTS | JUSTIFICATION FOR ENDPOINTS |
| --- | --- | --- |
| **Primary** |  |  |
| To determine whether iPS cells differentiated to airway epithelia -- and treated with ivacaftor/tezacaftor/elexacaftor *in vitro* -- can predict clinical responsiveness to ETI *in vivo*. | Functional and biological correction of CFTR expressed in iPS cells as judged by short circuit current measurements in monolayers, and western blot analysis. | These *in vitro* tests represent standard functional and biochemical analyses that address CFTR rescue, with direct relevance to the *in vivo* situation. |
| To evaluate FEV<sub>1</sub> clinical responsiveness to ETI *in vivo*. | Forced expiratory volume in one second (FEV<sub>1</sub>). | FEV<sub>1</sub> is a conventional endpoint and provides a direct measurement of patient health benefit among individuals with NCFB. |
| **Secondary** |  |  |
| To verify safety and further establish efficacy of ETI in enrolled patients. | Sweat chloride test<br>Quality of life survey<br>Standard laboratory tests and other measurements, as well as history and physical examination | Provide supportive information regarding study intervention effect using *in vivo* measures known to indicate CFTR rescue<br>A well-established tool for monitoring patient benefit<br>Confirm safety of the FDA-approved drug |

### Study Intervention/Investigational Agent

Subjects will be given the Study Drug elexacaftor 100 mg/tezacaftor 50 mg/ivacaftor 75 mg (2 pills once daily in the morning) and ivacaftor (150 mg) once daily in the evening, as the FDA-registered agent, ETI. Dose and schedule will be identical to what has already been FDA-approved for effective treatment of CF. The research pharmacy will be used since ETI is not on label for NCFB.

### Procedures Involved

#### Human Subjects Involvement

| Procedures | Screening/<br>Enrollment<br>Day -28 to -1 | Baseline<br>Visit 1, Day 1 | Study Visit 2<br>(phone call)<br>Day 7 +/-2<br>days | Study Visit 3<br>Day 14 +/- 2<br>days | Study Visit 4<br>Day 28 +/-2<br>days | Final Study<br>Visit Day 56<br>+/-2 days |
| --- | --- | --- | --- | --- | --- | --- |
| Informed consent | X |  |  |  |  |  |
| Medical history | X | X | X | X | X | X |
| Physical exam | X | X |  | X | X | X |
| Cutaneous punch biopsy and/or blood sample<sup>A</sup> |  | X |  |  |  | X<sup>D</sup> |
| CFTR genotype | X |  |  |  |  |  |
| Spirometry | X | X |  | X | X | X |
| Oxygen saturation | X | X |  | X | X | X |
| Sweat chloride testing | X | X |  | X | X | X |
| Safety laboratory assessment<sup>B</sup> | X | X |  | X | X | X |
| Pregnancy Test<sup>C</sup> | X | X |  |  |  | X |
| EKG | X |  |  | X |  |  |
| Drug accountability |  | X |  | X | X |  |
| Quality of Life (QOL) Questionnaire | X | X |  | X | X | X |
| Administer Study Drug |  | X |  |  | X |  |
| Concomitant medication | X | X | X | X | X | X |
| Adverse event assessment |  | X | X | X | X | X |
| <sup>A</sup> Blood sample or cutaneous punch biopsy for iPSC derivation and differentiation to airway epithelial monolayers<br><sup>B</sup> Renal and liver panels, CBC/Diff, urinalysis<br><sup>C</sup> Females of reproductive age or capable of becoming pregnant based on study physician evaluation<br><sup>D</sup> If necessary to produce additional iPS cells |  |  |  |  |  |  |

The study will examine iPSC-derived airway epithelial monolayers (an *in vitro* analytic tool) as a predictive means for identifying individuals with NCFB most likely to show respiratory benefit from ETI. These airway cells will be generated from blood and/or skin samples taken from participants. Enrollment is planned at Emory University. Patients in the trial will have one disease-causing CFTR variant and/or modestly elevated sweat chloride values (30-59 mEq/L), but without clinical findings sufficient for a diagnosis of cystic fibrosis.

Human subject participation in the ‘main’ study will consist of a screening day (−28 to −1), day 1 (+/-2), day 7 (+/-2) [phone call visit], day 14(+/-2), day 28 (+/-2) and day 56 (+/-2) follow up (wash-out). The visits will include patient history, physical exam, safety laboratory assessments (renal and liver panels, CBC/Diff, urinalysis), serum and urine pregnancy testing, CFTR genotype, spirometry, O_2_ saturation, sweat chloride test, blood sample collection and/or cutaneous punch biopsy for iPSC derivation, EKG, drug accountability and questionnaire for quality of life (QOL assessment), concomitant medication review, and adverse event assessment. Longer-term monitoring (termed “follow-on” study) for 12 months will be offered to patients who complete the “main study”. This will allow evaluation of any unexpected sequelae of ETI in the NCFB population (a group significantly older than most patients with CF). In addition, for individuals who exhibit favorable ETI response in the “main study”, durability of the benefit will be monitored. This will include clinical findings for patients who use data from the “main study” to obtain insurance reimbursement for ETI. The 12-month “follow-on” study will utilize a separate consent document.

#### “Main Study” Inclusion and Exclusion Criteria

We anticipate the screening and enrollment visit for the ‘main’ study will require 4-6 hours to complete the informed consent, medical history, O_2_ saturation, concomitant medication review, physical exam, safety laboratory assessments (CBC/Diff, RENAL/LFT chemistry panel, and urinalysis), EKG, CFTR genotyping, questionnaire, serum pregnancy test for females capable of pregnancy, sweat chloride test, and spirometry.

##### Inclusion Criteria

- Provision of signed and dated informed consent form
- Stated willingness to comply with all study procedures and availability for the duration of the study
- Male or Female age ≥18
- Radiologic and other clinical evidence leading to a diagnosis of NCFB
- 1 CF-causing mutation and/or sweat chloride measurement ≥ 30 mEq/L and < 60 mEq/L
- Able to perform spirometry meeting ATS criteria for acceptability and repeatability, and FEV_1_ 40-90 % predicted. Clinically stable in the past 4 weeks with no evidence of bronchiectasis exacerbation (prior to Screening AND Day 1).
- Willingness to use at least one form of acceptable birth control, including abstinence or condom with spermicide. This will include birth control for at least one month prior to screening and agreement to use such a method during study participation for an additional four weeks after the last administration of Study Drug. For postmenopausal or other women who are without the possibility of becoming pregnant, this requirement may be waived if a physician deems the study subject as “N/A”.
- Ability to take ETI
- Agreement to adhere to all current medical therapies as designated by the study physician

##### Exclusion Criteria

- Diagnosis of cystic fibrosis
- Documented history of drug or alcohol abuse within the last year
- Pulmonary exacerbation or changes in therapy for pulmonary disease in the 4 weeks prior to screening
- Listed for lung or liver transplant at the time of screening
- Cirrhosis or elevated liver transaminases > 3X upper limit of normal
- Pregnant or breastfeeding
- Inhibitors or inducers of CYP3A4, including certain herbal medications and grapefruit/grapefruit juice, or other medicines known to negatively influence ETI administration
- History of solid organ transplant
- Active therapy for non-tuberculosis mycobacterial infection or any plan to initiate non-tuberculosis mycobacterial therapies during the study period
- Known allergy to ETI
- Treatment in the last 6 months with an approved CFTR modulator
- Any other condition that, in the opinion of the lead investigators, might confound results of the study or pose an additional risk from administering Study Drug
- Treatment with another investigational drug or other intervention within one month prior to enrollment, throughout the duration of study participation, and for an additional four weeks following final drug administration.
- Evidence of cataract/lens opacity determined to be clinically significant by an ophthalmologist at or within 3 months prior to the Screening Visit

##### Expected Outcomes

Please note that based on past experience conducting clinical trials, we do not have concerns regarding our ability to perform the patient-oriented study described here, or to propagate and characterize iPS cells^19-21,26^. The ‘main’ study protocol will evaluate clinical benefit of ETI in patients with NCFB. In addition, the study will furnish one test of our hypothesis that thresholds of CFTR activity in differentiated iPS cells *in vitro* can predict benefit following ETI *in vivo*. If a strong correlation can be established between ETI response *in vitro* and *in vivo* (e.g., FEV_1_), the results will provide evidence regarding usefulness of *in vitro* models for predicting clinical efficacy in this disease setting.

#### Statistical Analysis Plan

Goals of the “main study” are to determine the clinical response to ETI and whether *in vitro* responsiveness of iPS cell-derived airway monolayers predict ETI effectiveness. Primary outcomes comprise the ability of iPSC-derived monolayers to predict respiratory benefit from ETI after 4 weeks of *in vivo* treatment. Primary analysis will include an estimate of clinical response rate of ETI at week 4 of treatment and a 95% confidence interval using the exact method proposed by Blaker^27^. A responder is defined as any subject with an improvement, from baseline, in FEV_1_ > 5% predicted. FEV_1_ will also be considered continuously. A two-sided 0.05 alpha level paired t-test comparing the baseline and week 4 FEV_1_ measurements, along with the test and 95% associated confidence interval, will be evaluated.

In this study, if at least 15% of subjects meet the definition of responder, we will view this as initial evidence of a favorable result. Overall response rate will be accompanied by an exact 95% confidence interval. To determine the role of iPS *in-vitro* response to ETI, we will compare the Δ lsc (change in lsc) among ETI responders and ETI non-responders using an ANOVA test. Additionally, we will correlate the Δ lsc obtained from *in vitro* experiments with the change in FEV_1_% predicted (post minus pre) using Spearman’s rank-order correlation with an associated 95% confidence interval. We will also test whether the iPS response reaches a CFTR functional threshold of 30% wild-type levels (a magnitude of activity in primary airway epithelial cells viewed as potentially relevant to clinical benefit). In a sensitivity analysis, we will replicate the FEV_1_ analysis with sweat chloride responses and determine the *in vivo* response rate.

Secondarily, we will serially evaluate clinical outcomes of interest prior to and after the start of therapy. Change in clinical measurements (in addition to spirometry – sweat chloride, weight/BMI, quality of life) will be monitored. A complementary analysis evaluating the percentage of participants with ≥15 mmol/L decline in sweat chloride will also be reported. With a previously estimated 9.7 mmol/L standard deviation of the change in sweat chloride for successful CF modulator trials^28,29^, an anticipated 6% of responders would be expected by chance alone. Therefore, the treatment response prevalence will be compared to a reference rate of 6%. For longitudinal FEV_1_ through day 28 and for secondary endpoints (absolute change in sweat chloride, QOL questionnaire, and weight/BMI from baseline through day 28), means at each time point and changes over time will be summarized along with corresponding confidence intervals estimated by mixed effects models to account for within-subject correlation.

Changes in FEV_1_ following treatment with ETI will be correlated with secondary clinical outcome measures. A non-linear Spearman’s correlation coefficient will be estimated for each measure paired with iPS cell responses *in vitro*, with corresponding fitted curves displayed graphically along with coordinates for participant’s paired *in vivo* and *in vitro* responses.

For other outcomes of interest including sweat chloride levels, quality of life, and weight/BMI, we will also apply a mixed effect model to examine the change in these measures over the study duration. Models will include a subject-specific intercept, to account for variability among subjects prior to the start of the treatment, and a categorical variable for time (baseline, day 14, day 28, day 56). A Dunnet’s post-hoc comparisons procedure will be used to determine which time points demonstrated a significant change from baseline. Using the same mixed effects models, we will include Δ Isc as predictors of *in vivo* improvement. We will also test the interaction between time and Δ lsc to determine when *in vitro* response is most associated with change in clinical response. Adverse events and other safety endpoints will be tabulated and reported using counts and percentages or means and standard deviations, as appropriate.

Because clinical parameters may vary among patients prior to the start of therapy, we will also approach the analysis using a modified N of 1 design configured for the purpose of comparing iPS studies in contrast to the aggregate analysis described above. Using the framework of single subject research, we will evaluate therapeutic response in individual subjects by an interrupted time series (ITS) method. Briefly, in the absence of an independent control group, ITS analysis is a quasi-experimental design in which each subject acts as their own control and is followed serially in time prior to and after an interruption (in this case, the start of treatment). If the treatment has causal impact, the post-intervention time series will have a different level or slope than the pre-intervention series. Time series models also have the power to test and correct for possible cyclical patterns and outliers. Using segmented regression analysis, we will conduct ITS models for each of the subjects enrolled in the trial. A significant change in slope from the pre-treatment period will indicate a significant treatment effect. ITS models will be constructed for each outcome of interest where historical and pre-treatment data are available. The change in slope will also be correlated with the *in-vitro* response using Spearman’s rank-order correlation coefficient.

All analyses will be conducted using the intention to treat principal, where all subjects will be included in analysis regardless of how long they took the study medication. Statistical significance will be assessed at the 0.05 level and analyses conducted using SAS v. 9.4 (SAS Institute; Cary, NC). (See also Supplemental Material S4)

## Data Availability

All data produced in the present study are available upon reasonable request to the authors.

## Notes

### Competing Interest Statement

The authors have declared no competing interest.

### Clinical Trial

NCT05743946

### Funding Statement

This study was funded by The Marcus Foundation, Inc., Project #00117885. No other third-party payment or services were received by authors or their institutions for any aspect of the submitted work. The Marcus Foundation had no involvement in research design or decision to publish the attached protocol.

### Author Declarations

Ethics committee/IRB of Emory University School of Medicine gave ethical approval for this work.

