## Supplemental Materials S1-S4 for "Evaluating elexacaftor/tezacaftor/ivacaftor (ETI; Trikafta™) for treatment of patients with non-cystic fibrosis bronchiectasis (NCFB): a clinical study protocol"

**Table of Contents** **Page**


| | |
|---|---|
| **S1 – Details regarding study activities** | **2** |
| **S2 – Risk to Participants** | **4** |
| **S3 – Scientific and clinical support for the clinical trial** | **6** |
| **S4 – Additional Statistical Considerations** | **12** |
| **References** | **14** |

**S1 – Details regarding study activities**

Sources of Patient-Derived Material for "Main Study"

- Review of patient medical records and demographic information
- History and physical exam
- Blood and urine for safety studies (<6 tablespoons of blood per visit)
- Sweat chloride results
- Questionnaire data
- Spirometry ($FEV_1$, FVC, and FEF25-75%)
- Cutaneous punch biopsy (3 mm) and/or blood sample (8 ml) for iPS cell derivation
- $O_2$ saturation
- EKG
- CFTR genotype (evaluated by the study or collected from the patient's medical record)
- Pregnancy test (female capable of becoming pregnant, waived at physician discretion)
- Drug accountability
- Adverse event assessment
- Concomitant medication review

The "follow-on" study will include a separate consent and comprise 5 clinic visits – at entry (day 0), 3 months, 6 months, 9 months, and 12 months – after signing a "follow-on" trial consent. Testing at each visit will be identical to the day 56 ("wash-out") visit in the "main study," except that pregnancy test (if deemed necessary by study physician) will occur only at the day 0 visit, no skin biopsy or blood samples for iPSCs will be obtained, and pulmonary CT scan will be performed during the day 0 and at 6-month and 12-month visits.

Subject recruitment plan

For the lead-in study, patients followed at Emory with a diagnosis of NCFB will be sent an IRB-approved email from their physician one to two weeks before their next pulmonary visit. If there is no response to the email (or if favorable response to the email), patients will be contacted by phone before the clinic visit and offered a chance to review the lead-in study informed consent document. Informed consent will be discussed and possibly signed or declined at the clinic visit.

For the main study, we plan to recruit 30 subjects with NCFB. We intend to include ~50% female and ~50% male subjects age ≥ 18. Subjects will be referred from clinicians participating in the trial (Emory University, Atlanta) or outside institutions.

All individuals who complete the main study will be offered an opportunity to participate in the follow-on study.

1. Informed Consent Process:

Informed consent will be obtained by well-trained study coordinators or investigators using an Institutional Review Board-approved document. A consent to allow CFTR genotyping and sweat chloride will be offered as part of the 'lead in' study to identify potential subjects with CFTR mutations or mildly elevated sweat chloride levels. A second consent form will be available regarding the "main study" for individuals found to have a single disease-causing CFTR mutation and/or sweat chloride 30-59 mEq/L, but without clinical criteria necessary for a diagnosis of cystic fibrosis. Participants will be asked to sign the informed consent documents only after the study has been fully described and all questions have been answered. Participants will be offered at least 24 hours to review the consent before the research is to begin, although consent may be obtained on the day of screening if preferred by study subjects (e.g., those traveling from a significant distance). In certain instances, it may be possible for patients who possess prior knowledge regarding their mutation and/or sweat chloride levels to potentially skip the “lead-in” study and proceed straight to the “main study”. This option may be available to those who meet specific criteria and have been approved by their medical team. It is important to note that this decision should only be made after careful consideration by the healthcare professionals involved in the study.

NCFB is not associated with cognitive or other intellectual limitations. No vulnerable adult subjects are intended for enrollment in the trial. If, in the opinion of the study investigator, a prospective participant lacks sufficient understanding and is therefore not able to provide informed consent, that subject will not be enrolled. The ability to obtain consent represents criteria for inclusion in the trial.

### S2 – Risk to Participants

**Blood Draws**
Possible risks include hematoma at the needle site and minimal pain due to the venous puncture procedure.

**Sweat chloride testing**
The Macroduct system is used in hospitals and clinics worldwide to perform sweat chloride testing. This is considered a very safe, painless procedure, but rare minor burns have been reported at the electrode site (although participants may show no discomfort during the test). To further minimize risk, all equipment will be properly cleaned and evaluated before and after the procedure by personnel trained in performance of this assay.

**Risks of the punch biopsy**
Significant pain, bleeding, or infection are very rare with this type of punch biopsy. Investigators may use a topical anesthetic on the skin in order to minimize pain, although study subjects might feel minor discomfort (like a scratch) when the topical anesthetic is administered, and the next day. There is the possibility of a small scar.

**Risks from Study Drug (elexacaftor 100 mg/tezacaftor 50 mg/ivacaftor 75 mg (2 pills once daily in the morning) and ivacaftor (150 mg) once daily in the evening)**
There may be side effects from the Study Drug or procedures that are not known at this time. Examples of adverse reactions known in patients receiving ETI are listed below.

Some of the most common (occurring 5% or more) risks and discomforts expected in this study are:

| | |
|---|---|
| Headache | 17% |
| Upper respiratory tract infection | 16% |
| Abdominal pain | 14% |
| Diarrhea | 13% |
| Rash | 10% |
| Blood ALT increased (liver marker) | 10% |
| Nasal congestion | 9% |
| Blood CPK increased (muscle marker) | 9% |
| Rhinorrhea (runny nose) | 8% |
| Rhinitis (nose irritation) | 7% |
| Blood AST increased (liver marker) | 9% |
| Influenza | 7% |
| Sinusitis | 5% |
| Blood bilirubin increased (liver marker) | 5% |

Increased liver enzymes (ALT or AST) in the blood have been observed in some subjects. Very high levels of these enzymes could lead to stopping of Study Drug. The abnormal blood tests may improve after Study Drug is stopped. In some severe cases, high liver enzymes can become permanent or even life-threatening.

Possible Risks Based on Animal Studies

In a study in which ivacaftor (a component of ETI) was given to newborn rats, cataracts (cloudiness of the lens of the eye) were seen. No cataracts were seen in studies of older animals (rats and dogs) dosed with ivacaftor for longer periods of time. The importance of this finding in humans is unknown.

Drug Interaction Risks

The combination of Study Drug and other medications, dietary supplements, natural remedies, and vitamins could be harmful to trial participants. Subjects will be asked to inform study staff about every medicine, dietary supplement, natural remedy, and vitamin (or change in medicine) while they are in the study. The study staff will review all medications (including herbal medications, such as St. John's Wort, grapefruit or grapefruit juice) that should not be utilized during the study because herbal compounds such as these can alter metabolism of ETI.

Risks of Discontinuing Study Medication:

Once the four-week treatment period is complete, participants will no longer receive ETI as part of the trial. There is a chance that the NCFB will improve when taking ETI. However, NCFB symptoms may appear to worsen after the participant stops taking ETI as respiratory function returns to baseline.

Women:

To protect against possible side effects of Study Drug, women who are pregnant or nursing a child may not take part in this study. If a participant becomes pregnant, there may be risks to the participant, the embryo, or fetus. These risks are not yet known. If the participant is a woman of childbearing ability, the participant and the Study Doctor must agree on a method of birth control to use throughout the study. Pregnant women will be taken out of the study.

Possible risks to non-study participants:

The effect of the Study Drug on sperm is not known. To protect against possible side effects, if the female partner of a male participant becomes pregnant during the Study, they should notify the Study Doctor immediately.

Study Drug should be kept out of the reach of children or anyone else who may not be able to read or understand the label. Participants should not allow anyone else take the Study Drug.

It is possible that new information regarding Study Drug or participation in the trial during the course of the clinical protocol will become evident. If this happens, patients will be informed so that they can make a decision about continuing in the study. Participants in the trial may be asked to sign a new consent form that includes the new information in order to continue in the study.

Potential Benefits to Participants

This study is not designed to benefit the participant directly. NCFB may improve in this study but it may not, and it may also become worse. This study will provide new information regarding whether cells from the skin or blood can be used to predict patients with NCFB who respond best to ETI.

### S3 – Scientific and clinical support for the clinical trial

#### A. Background and Rationale: NCFB and relationship to cystic fibrosis

Like NCFB, cystic fibrosis (CF) bronchiectasis is associated with structural dilatation of the airways, mucus stasis, chronic bacterial infection, lung tissue remodeling, and diminished respiratory function[1-6]. CF is an autosomal recessive illness attributable to absence or dysregulation of the cystic fibrosis transmembrane conductance regulator (CFTR), an epithelial ion channel situated within apical membranes of secretory epithelium and essential for mucociliary clearance by the lung.

Highly effective CFTR modulator treatments (HEMTs) such as elexacaftor/tezacaftor/ivacaftor (ETI) have been shown to provide dramatic benefit among patients with CF[6-11]. Modulators work by augmenting biogenesis, maturation, and/or gating of the CFTR ion channel. Based on an increasing body of evidence, the current clinical trial will test whether certain patients with otherwise poorly treatable NCFB might demonstrate clinical improvement from compounds that activate CFTR. That notion has not been adequately proposed or evaluated, although scientific and mechanistic rationale are strong.

As a first step, we are investigating whether modulators that activate both wild-type and mutant CFTR will improve NCFB clinical status, and whether clinical findings can be predicted using an iPSC-based model system.

#### B. Epidemiologic and clinical findings in support of the trial

B1. Data showing CFTR deficiency exists along a continuum in patients with bronchiectasis
One can argue that discriminating between clinically important subtypes of bronchiectasis might be improved by considering the extent to which CFTR is functionally available. For many patients with a diagnosis of bronchiectasis, both sweat chloride and CFTR ion channel activity (using cell systems relevant to FDA modulator approval) are known to distribute across a spectrum (**Figure A**)[12]. Earlier reports indicate substantial numbers of patients with NCFB exhibit diminished CFTR function[13-18], and provide impetus for the current clinical study. Patients with NCFB who are deficient for CFTR might benefit from ETI (which robustly activates both mutant and wild-type CFTR).

B2. Diagnostic considerations relevant to the trial
Testing for CFTR reserve is not typically performed as part of standard management for patients with NCFB. If a person with bronchiectasis is older and lacks extrapulmonary manifestations of cystic fibrosis, the diagnosis of NCFB is commonly applied. If the present clinical trial is successful, the findings will suggest that diagnostic criteria for NCFB can be enhanced by expanded monitoring for CFTR deficiency (e.g., as judged by mutation analysis, iPSC measurements, elevated sweat chloride, and/or bioelectric studies *in vivo*). The latter interpretation has already been acknowledged for individuals who exhibit an incomplete CF phenotype (i.e., CFTR-related disorder[19]). Importantly, among patients with "CFTR-related disorder", a single F508del mutation does not allow access to CFTR modulators. The drug is costly—and "off label" use (for patients without a formal diagnosis of cystic fibrosis)—is not typically reimbursable by insurers. That said, individuals with CFTR-related disorder commonly exhibit evidence of both CFTR deficiency and bronchiectasis, and there is every reason to believe that in many such patients, respiratory improvement would result from ETI. From the standpoint of a much larger patient population, the label of

“non-CF bronchiectasis” is often assigned (in general medical and pulmonary clinics worldwide) without evaluating either CFTR genotype or sweat chloride, and a diagnosis of NCFB largely precludes treatments such as ETI. This is despite the fact that a genotype encoding a single CFTR mutation such as F508del is more common in patients with NCFB and is clearly associated with pathogenesis[13-15,17,18]. Although mechanistic overlap surely exists between many cases of CFTR-related disorder and NCFB, ETI has never been evaluated in controlled studies for either condition. Rigorous trials such as the one described here are needed to investigate CFTR modulators in patients with NCFB.

#### C. Caveats regarding ETI as a potential intervention for NCFB

Medical clinics internationally follow large numbers of individuals with NCFB diagnosed and managed in the same manner as those intended for the current study—but without consideration of CFTR genotype testing or treatment with ETI. This traditional approach to NCFB diagnosis and management has been acceptable in part because interventions for addressing CFTR deficiency were not available, and determination of adequate CFTR function therefore was not pursued. With the advent of highly effective modulator treatments (and if the present clinical trial confirms our hypotheses regarding predictive value of iPSCs for patient benefit), we suggest a standard NCFB diagnosis would be aided by including an assessment of CFTR reserve.

#### D. Relevance of drug cost

The high price of CFTR modulators prevents ETI access globally among patients with NCFB, CFTR-related disorder, and many patients with CF. While modulator treatment is well tolerated in most individuals with cystic fibrosis (as well as the NCFB patients anecdotally described below), formal (and very costly) clinical trials[20,21] in much larger numbers of subjects will be needed to evaluate safety and efficacy among those with NCFB, and to determine the extent to which this patient population might benefit. At least in principle, if our trial is successful, this could impact the high cost of ETI therapy by significantly expanding market size.

#### E. Examples of clinical experience using ETI treatment for NCFB

Our research team has evaluated some of the first patients in the world with NCFB who have been treated using ETI. As one example, a female in her 60’s with a diagnosis of NCFB and refractory nontuberculous mycobacterial lung disease (NTM-LD) was referred for evaluation. Over a decade earlier, the patient completed a prolonged course of daily ethambutol, rifampin and azithromycin for treatment of *M. avium* complex (MAC). Sputum subsequently grew *M. abscessus* complex, and clofazimine, amikacin, tigecycline, and imipenem-cilastatin were administered, followed by clofazimine, azithromycin, bedaquiline, and nebulized amikacin. The patient subsequently experienced significant weight loss, daily low-grade fevers, and intermittent hemoptysis. CT imaging demonstrated NCFB (**Figure B, Panel A**). During follow-up, further weight loss and productive cough were noted. In addition to positive mycobacterial cultures, sputum grew *Stenotrophomonas* and *Pseudomonas aeruginosa.* At presentation to our clinic, and in an attempt to eradicate *P. aeruginosa*, levofloxacin followed by nebulized tobramycin were initiated. While sputum production and hemoptysis improved, weight loss, fatigue, and low- grade fever persisted. COPD Assessment Test (CAT) score was 27, indicating a considerable symptom burden. Forced expiratory volume in 1 second (FEV1) and body mass index (BMI) were significantly decreased (**Table 1**). Immunoglobulins, alpha 1 antitrypsin level, and autoimmune testing were unrevealing, whereas CFTR genotype demonstrated a single copy of F508del following complete CFTR DNA sequencing.

In the setting of declining clinical status, refractory NTM, and ongoing infections with gram-negative pathogens, and based on F508del carrier status, ETI was initiated. At follow-up visit several months later, the patient reported near total resolution of her prior respiratory and constitution symptoms. She retained mild intermittent cough without significant sputum production. There was no further hemoptysis, fever, or weight loss, and BMI increased. CAT score was 10, reflecting a substantial response of symptomatology. CT imaging revealed less mucus impaction, diminished tree-in-bud nodularity, and decreased air trapping (**Figure B, Panel B**). The pronounced symptomatic and clinical benefit have persisted beyond one year while continuing ETI, with considerable improvement of FEV1.

As another example, a female in her 60's with a chronic history of NCFB and sputum positivity for MAC and *Pseudomonas aeruginosa* had been treated with clarithromycin, ethambutol, and moxifloxacin. Sputum became negative for MAC, but continued to grow *P. aeruginosa*, as well as *Aspergilus fumigatus*, *Scedosporium apiospermum*, and methicillin sensitive *Staphyloccus aureus* (MSSA). Oral antibiotics (e.g., fluoroquinolones) were administered intermittently for many years, together with two courses of itraconazole. Upon transfer to our clinic, the patient reported daily cough productive of green sputum and hemoptysis. She also complained of fatigue, dyspnea on exertion, and recurrent low-grade fever. CAT score was 25, reflecting high disease burden. Laboratory evaluation revealed a single copy of F508del CFTR (from a screen for 165 CFTR variants), and respiratory culture yielded two isolates of mucoid *P. aeruginosa* (fluoroquinolone resistant). The patient was prescribed cycled nebulized tobramycin, but CAT scores remained elevated, with respiratory samples that continued to grow *P. aeruginosa*, MSSA, and *Aspergillus* species. MAC was consistently isolated from surveillance respiratory cultures, with worsening of clinical symptoms and diagnostic radiographic findings (**Figure B, Panel C**). Treatment was initiated with a combination of daily azithromycin, ethambutol and clofazimine. Fever resolved and cough improved, but fatigue, weight loss, and intermittent hemoptysis persisted.

Based on severe pulmonary insufficiency and debilitating infection despite aggressive antimicrobial therapy, the patient was prescribed ETI. At follow-up several weeks later, all symptoms had markedly improved, with CAT score falling to 9. Spirometry and BMI increased (**Table 1**), and radiographic improvement was noted (**Figure B, Panel D**). These favorable findings have been maintained for over a year on ETI.

A third vignette involves a male in his 50's presenting with a near decade-long history of NCFB. Cultures were positive for methicillin-resistant *staphylococcus aureus* (MRSA) and *Mycobacterium abscessus massiliense*. CT scan demonstrated moderate cylindrical bronchiectasis and tree-in-bud infiltrates affecting both upper lobes, right middle lobe and superior segments of the right and left lower lobes. Spirometry demonstrated an FEV1 of 2.92 L (89% of predicted). Complete CFTR DNA sequencing revealed a single copy of F508del. Sweat chloride levels were 25-28 mEq/L, and fecal elastase was within normal limits. The patient was started on airway clearance therapy with a flutter valve and hypertonic saline, but experienced worsening of cough, and sputum cultures remained positive for MRSA. Treatment with doxycycline conferred minimal benefit. Repeat chest CT demonstrated NCFB progression with new areas of nodularity and airway impaction in the left upper and right middle lobes. Sputum grew *Mycobacterium abscessus* complex and FEV1 continued to decrease. Anti-mycobacterial treatment led to improvements in cough, fatigue, and dyspnea, with sputum negative for acid-fast bacilli (AFB). The patient remained on antimicrobials, but cultures were found to contain *Pseudomonas aeruginosa, Pseudomonas putida*, *Stenotrophomonas* and *Aspergillus*, together with reappearance of *Mycobacterium abscessus* complex. This coincided with increased cough and sputum production.

A trial of ETI was undertaken when spirometry (FEV1 2.36 L; 76% predicted), BMI, and clinical trajectory were deteriorating (**Table 1**). Shortly after starting CFTR modulator therapy, the patient reported improved breathing, diminished cough, and thick yellow sputum had become scant. Fatigue diminished and exertional capacity increased. Spirometry and BMI improved (**Table 1**). Bronchiectasis Severity Index (BSI) scores improved from 8 to 3. Subsequent sputum cultures (while on ETI) have remained negative for MRSA, mycobacterium, and *Pseudomonas* for over a year, with respiratory symptoms continuing to benefit and FEV1 remaining stable.

#### F. Strong rationale for the clinical trial

Hundreds of thousands of individuals with NCFB worldwide who receive standard medical care have not been evaluated for evidence of partial CFTR deficiency. Based on emerging knowledge of CFTR biology, airway physiology, population genetic studies, clinical epidemiology, and other scientific evidence, we believe many patients with NCFB are likely to benefit from CFTR activation and the associated increase in mucociliary clearance. Modulator responsiveness for NCFB has not been adequately considered, in part because the two diseases (NCFB and CF) have been viewed as mechanistically distinct and mutually exclusive. Clinical trials such as the one described here will help determine whether patients with NCFB can experience lung function improvement using ETI and whether iPSC derived airway epithelial monolayers can identify individuals with NCFB most likely to exhibit therapeutic benefit. A refined mechanistic understanding of the disease is also likely to result (**Figure C**). If the hypotheses pursued here are validated by the current clinical protocol, the data will offer new therapeutic options for patients with an otherwise poorly treatable and potentially lethal lung disease.

**Table 1: FEV1 (in L(%predicted)) and BMI (kg/m$^2$) before and after ETI Treatment**

| Case | Sex | FEV1 Pre-ETI | FEV1 Post-ETI | BMI Pre-ETI | BMI Post-ETI |
|---|---|---|---|---|---|
| 1 | F | 1.64 (66%) | 1.86 (76%) | 17.8 | 21.3 |
| 2 | F | 0.65 (29%) | 0.82(35%) | 18.8 | 19.9 |
| 3 | M | 2.36 (76%) | 2.45 (81%) | 24.6 | 25.1 |

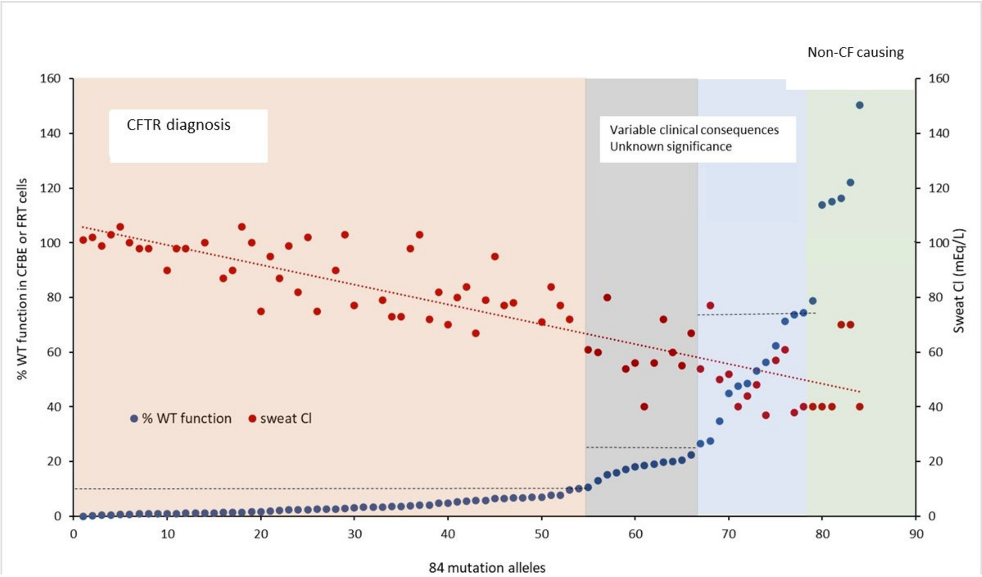


**Figure A. CF-related lung disease in relation to CFTR reserve.** Clinical data shown is from the CFTR2 database (https://cftr2.org/) or cell models reported by Han *et al*.[12]. Findings describe 84 CFTR mutations ranked according to *in vitro* activity. Sweat chloride values are based on patients in which the second CFTR variant encodes a mutation with minimal function. In that scenario, sweat chloride of 30-59 mEq/L is often associated with bronchiectasis and can be viewed as a risk factor for CFTR-deficient lung disease. “Variable clinical consequences/Unknown significance” includes patients with bronchiectasis but clinical findings inadequate to establish a complete CF phenotype. FRT: Fischer rat thyroid cells. CFBE: cystic fibrosis bronchial epithelium. Horizontal dashed lines represent upper threshold of CFTR activity in relation to wild-type CFTR in cell lines for each diagnostic group.

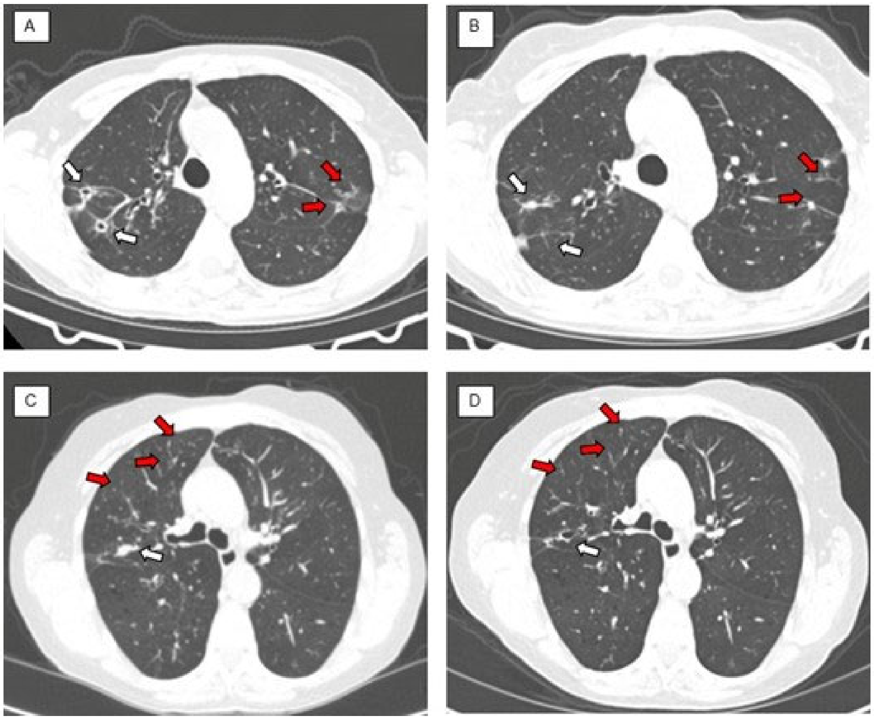


**Figure B. CT chest images for patients with NCFB described in text. (A)** CT chest image pre-ETI demonstrates cavitary lesions in right upper lobe (white arrows) and inflammatory changes in left upper lobe (red arrows). **(B)** CT chest image post-ETI showing closure of cavitary lesions (white arrows) and cicatricial healing in left upper lobe (red arrows). **(C)** Axial CT image before ETI. Note mucus impaction (white arrow) and tree-in-bud nodularity in the right upper lobe (red arrows). (**D**) After ETI, improvement in mucus impaction was observed (white arrow) together with resolution of tree-in-bud nodularity (red arrows).

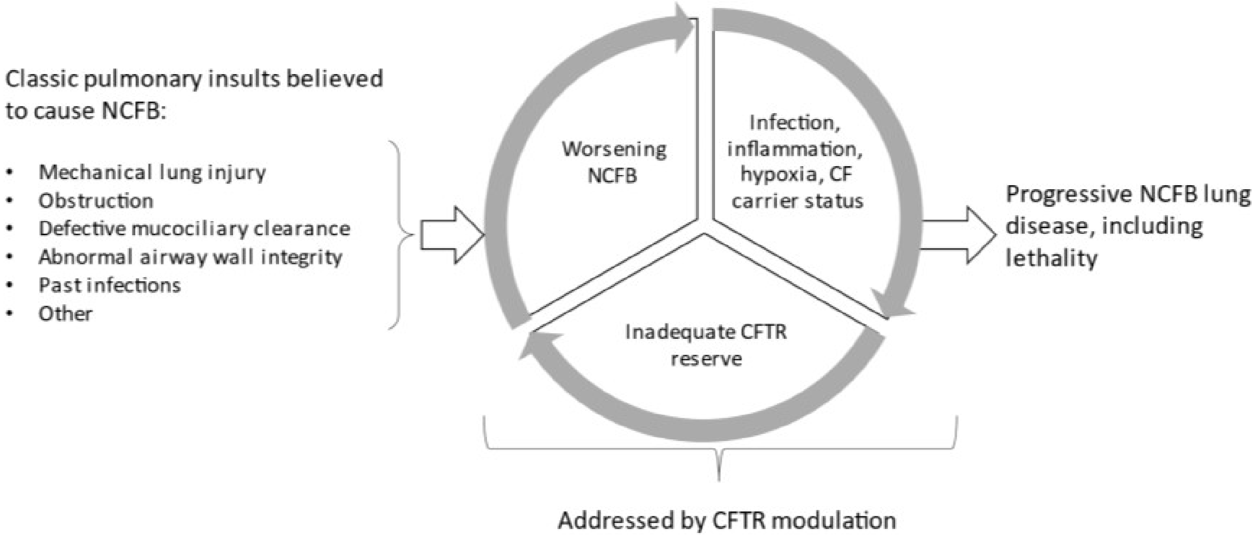


**Figure C. Mechanism of NCFB pathogenesis.**

### S4 – Additional Statistical Considerations

**Sample size**

We will test the null hypothesis that the true ETI response rate of NCFB individuals is ≤ 1% vs. the alternative hypothesis that the true ETI response rate is ≥ 15%. We will initially enroll 16 subjects. If none of these subjects exhibit a clinical response (defined as > 5% improvement of % predicted $FEV_1$ from baseline), the study may be considered for termination. Otherwise, we intend to enroll a total sample size of 29 individuals (or 30 subjects, in the unlikely event that an individual discontinues participation; i.e., subjects who do not complete the study will be replaced). In this design, if ETI is ineffective, the probability of early termination after 16 subjects is 85% and only a 2.8% chance of concluding the drug is effective (type I error). If ETI is effective, there is a 90.04% chance of reaching this conclusion. If fewer than 2 patients (of 29 patients total) of the overall cohort show a clinical effect, ETI will be considered ineffective. To demonstrate the ability of iPS cells to predict clinical benefit, 16 (29) patients will provide at least 80% power to detect a significant Spearman correlation of at least r = 0.8 (.53) between the changes in clinical outcomes with changes in short circuit current as measured in iPS cells. Tabular details of this analysis are provided below. Power calculations were performed using PASS v. 14 (Kaysville, UT) with a two-sided Z-test and a significance level of 0.05.

| | **Cumulative # of responses** | **Decision** | **95% Confidence Interval (CI)** |
|---|---|---|---|
| **Stage 1: Enter 16 subjects** | 0 | Consider Termination of the trial because the agent is ineffective. **Response rate is ≤1%** | LB of 95% CI is < 1% |
| | At least 1 | Inconclusive result, continue trial (proceed to stage 2). | 95% CI contains 1% |
| **Stage 2: Enter 14 additional subjects** | 1 or less (stage I + stage II) | Consider that the agent is ineffective. Response rate is ≤1% | LB of 95% CI is less than 1% |
| | 2 or more (stage I + stage II) | Drug may be effective Response rate is greater than 1% and not different from 15% | 95% CI contains 15% and LB is > 1% |
| | 6 or more (stage I + stage II) | Drug is effective Response rate is at least 15% | 95% CI LB ≥ 15% |
| LB = Lower Bound, UB = Upper Bound<br>Note: Thirty total patients will be enrolled in the unlikely event that a subject discontinues participation (e.g., due to elevated liver function tests, skin rash, or for other reasons). | | | |

With a sample size of 16 (29) participants, we anticipate a 97% (>99%) power to detect a mean sweat chloride decrease of 10 mmol/L assuming a standard deviation of 9.7 mmol/L [22,23] under a two-sided 0.05 alpha-level paired t-test. With a higher standard deviation of 15 mmol/L, the power decreases to 70.3% (93.4%) to detect a mean decrease of 10, but we retain 96.2% (>99%) power to detect the hypothesized mean decrease of 15 mmol/L with a sample size of 16 (29) patients.

SAEs and AEs will be tabulated using standard coding terms sorted by System Organ Class (SOC). The incidence of AEs will be tabulated by seriousness and severity. The number of SAEs and AEs will be summarized as follows: (i) The proportion of subjects with at least one (S)AE, (ii) The average number of (S)AEs per subject, and (iii) The rate of (S)AEs per subject week of follow-up.
